# Assessment of twenty-two SARS-CoV-2 rapid antigen tests against SARS-CoV-2: A laboratory evaluation study

**DOI:** 10.1101/2021.12.15.21267691

**Authors:** Joshua M Deerain, Thomas Tran, Mitch Batty, Yano Yoga, Julian Druce, Charlene Mackenzie, George Taiaroa, Mona Taouk, Socheata Chea, Bowen Zhang, Jacqueline Prestedge, Marilyn Ninan, Kylie Carville, James Fielding, Mike Catton, Deborah A Williamson

## Abstract

**Background:** Rapid antigen testing is widely used as a way of scaling up population-level testing. To better inform antigen test deployment in Australia, we evaluated 22 commercially available antigen tests against the currently circulating delta variant, including an assessment of culture infectivity.

**Methods:** Analytical sensitivity was evaluated against SARS-CoV-2 B.1.617.2 (Delta), reported as TCID_50_/mL, cycle threshold (Ct) and viral load (RNA copies/mL). Specificity was assessed against non-SARS-CoV-2 viruses. Clinical sensitivity and correlation with cell culture infectivity was assessed using the Abbott PanBio™ COVID-19 Ag test.

**Results:** Nineteen kits consistently detected SARS-CoV-2 antigen equivalent to 1.3 × 10^6^ copies/mL (5.8 × 10^3^ TCID_50_ /mL). Specificity for all kits was 100%. Compared to RT-PCR the Abbott PanBio™ COVID-19 Ag test was 52.6% (95% CI, 41.6% to 63.3%) concordant, with a 50% detection probability for infectious cell culture at 5.9 log_10_ RNA copies/mL (95% CI, 5.3 to 6.5 log_10_ copies/mL). Antigen test concordance was 97.6% (95% CI, 86.3% to 100.0%) compared to cell culture positivity.

**Conclusions:** Antigen test positivity correlated with positive viral culture, suggesting antigen test results may determine SARS-CoV-2 transmission risk. Analytical sensitivity varied considerably between kits highlighting the need for ongoing systematic post-market evaluation to inform test selection and deployment.

## INTRODUCTION

The global COVID-19 pandemic has profoundly altered the way that societies live and function. To date, a combination of vaccination, local and national lockdowns, and contact tracing (‘test, trace, isolate and quarantine’) has been used to mitigate the transmission of SARS-CoV-2, with reverse-transcriptase polymerase chain reaction (RT-PCR) testing forming the cornerstone of testing in many countries (1, 2). In addition to molecular detection of SARS-CoV-2, point-of-care rapid antigen tests have been used to complement laboratory-based PCR testing. Deployment of rapid antigen testing has been suggested as a potential means of upscaling population-level testing to facilitate the safe re-opening of societies, and to enable access to COVID-19 testing in resource-poor settings (3-6). In theory, large-scale testing of asymptomatic populations using rapid antigen tests (in conjunction with other public health measures) could detect infectious individuals with pre-symptomatic or asymptomatic COVID-19 and rapidly interrupt transmission networks and is a conceptually attractive option for facilitating a ‘COVID Normal’ return to daily activities.

Because COVID-19 antigen tests detect viral protein (usually nucleocapsid) rather than amplified nucleic acid, they are inherently less sensitive than RT-PCR assays (7). However, their advantages include rapid results, portability and reduced cost compared to RT-PCR. Further, recent studies suggest that detection of viral antigen may correlate with higher viral load and hence, a greater likelihood of cell culture infectivity (8, 9). Although higher viral load does not necessitate a greater probability of infectiousness, regular antigen testing may serve to identify high risk individuals who are infectious and should be quarantined (10).

Australia has one of the highest per capita SARS-CoV-2 RT-PCR testing rates in the world; these high rates of testing have, along with major public health interventions, contributed to a relatively low incidence of COVID-19 (11). However, the recent emergence of the B.1.617.2 lineage (Delta lineage; World Health Organisation [WHO] nomenclature) in Australia in early 2021 has led to extensive community transmission in the two most populous states of Australia; New South Wales (NSW) and Victoria. These outbreaks have led to renewed interest in the application of antigen tests, with over 40 point of care lateral flow devices (LFD) assays now approved for use in Australia (as of 22^nd^ October, 2021 when this study commenced) by the Therapeutics Goods Administration (TGA), including two assays using saliva as the testing sample (12). Knowledge of the performance characteristics of antigen tests is critical in considering if, when and how they are most appropriately used. To date however, most published studies have not assessed a broad range of antigen test kits, and there are limited data available on the performance of antigen tests against the Delta variant. Accordingly, to better inform the deployment of antigen tests in Australia, we compared the performance characteristics of 22 commercially available antigen test assays, including an assessment of culture infectivity.

## METHODS

### Study samples and design

A laboratory evaluation study was conducted at the Victorian Infectious Diseases Reference Laboratory (VIDRL) at The Peter Doherty Institute for Infection and Immunity (Melbourne, Australia). VIDRL is the public health virology laboratory for the state of Victoria, serving a population of approximately 6.24 million people. The study was designed in two parts: (i) an evaluation of analytical sensitivity and specificity of 22 antigen tests, including two saliva antigen tests and two fluorescent immune assays (Supplementary Table 1); and (ii) an evaluation of clinical sensitivity and specificity of a high-performing antigen test and correlation with infectivity in cell culture.

### Analytical sensitivity and specificity

To evaluate analytical sensitivity and specificity, we used a dilution series of a representative isolate of the Delta variant currently circulating in Victoria (VIC/18440/2021). SARS-CoV-2 lineage classification was based on previously described methods (13). Virus was grown in Calu-3 cells (Supplementary Appendix) and inactivated by a 50kGy dose of gamma radiation. Gamma irradiation is commonly used in virology to inactivate viruses for subsequent use in the development and evaluation of laboratory assays, whilst preserving the structural integrity of surface antigens (14). To confirm that gamma irradiation had no impact on antigen kit evaluation, we compared the analytical sensitivity of an antigen test kit pre- and post-irradiation using VIC/18440/2021. Viral load was quantified by an in-house assay for the N-gene, described previously (15), and by droplet digital PCR (ddPCR; BioRad QX200) (Supplementary Appendix). Infectious virus was also quantified prior to gamma-irradiation by TCID_50_ assay (Supplementary Appendix).

Two-fold serial dilutions of quantified virus were prepared in either viral transport media (VTM) to simulate an eluted nasopharyngeal swab, or pooled healthy donor saliva, (that tested negative to SARS-CoV-2) ranging from approximately 10^1^ to 10^5^ TCID_50_/mL, covering a range of cycle threshold (Ct) values between 21-35 (based on the in-house N-gene RT-PCR) (Supplementary Table 2). Based on manufacturer’s reported sensitivities for assessed antigen test kits, we estimated that the limit of detection (LoD) for all devices would fall between 3 × 10^1^ and 2 × 10^3^ TCID_50_/mL. Samples with a Ct value between 21 and 23 (2 × 10^4^ and 1 × 10^5^ TCID_50_/mL, and > 33 (≥2 × 10^1^ TCID_50_/mL) were assessed in duplicate, while samples with a Ct between 24 and 32 were assessed in quadruplicate.

For the twenty nasal/nasopharyngeal antigen tests, testing was performed according to the manufacturers provided instruction for use (IFU) for assessing swabs eluted in VTM. Where no specific protocol was provided, a 1:1 dilution of sample to extraction buffer was prepared and added to the test kit using provided plasticware, as per manufacturer’s instructions (Supplementary Table 1).

To assess analytical specificity, panels of ‘distractor’ viruses commonly found in respiratory samples were prepared using ten stored reference isolates from VIDRL: Influenza A/B, Respiratory Syncytial Virus (RSV), Rhinovirus, human coronaviruses (OC43 and 229E), Adenovirus 3, Herpes Simplex Virus 1 (HSV1), Cytomegalovirus (CMV) and Parainfluenza virus 3. Samples were diluted in VTM or pooled saliva to a Ct ≤25 and quantified using ddPCR (Supplementary Appendix and Supplementary Table 3). Each sample was tested in duplicate (n=20 samples) using the same procedure as the sensitivity panels described above.

Testing duration and interpretation of antigen test results were performed as per manufacturer’s instructions. Results of all tests were recorded independently by two scientists. Results were read by a third independent scientist where there was discordance between initial readings.

### Assessment of clinical sensitivity and correlation with viral culture infectivity

Clinical sensitivity and correlation with cell culture infectivity was assessed using the Abbott PanBio ™ COVID-19 Ag test (Abbott Rapid Diagnostics Jena GmbH, Germany). This device was specifically chosen because clinical studies using this kit are ongoing in our setting. Residual VTM from combined naso-oropharyngeal swabs that were submitted to VIDRL for SARS-CoV-2 RT-PCR testing between 01/07/2021 and 22/08/2021 were stored at 4°C up to 1 week prior to testing. Samples were not subjected to a freeze-thaw cycle to facilitate preservation of virus for cell culture infectivity assessment. All samples reported as detected for SARS-CoV-2 and with sufficient volume remaining were included and assessed for the following: (i) antigen test reactivity; (ii) cell culture infectivity, and (iii) viral RNA load quantification using a quantitative standard curve (Supplementary Appendix). Lineage designation of SARS-CoV-2 was performed using previously described methods (13). Clinical specificity of the LFD was assessed using residual VTM from 100 SARS-CoV-2 RT-PCR negative combined naso-oropharyngeal swabs.

SARS-CoV-2 RT-PCR positive clinical samples were assessed for antigen test reactivity using the Abbott PanBio™ COVID-19 Ag Rapid Test Device (Nasal) (Abbott 41FK11). Samples were prepared and tested according to methods described in the ‘Guidance For Use of Alternative Protocol’ documentation provided by Abbott. Samples were diluted 1:1 in 150 µL extraction buffer and 130µL of the mixed solution added to the rapid test device. Results were recorded between 15-20 minutes according to manufacturer’s IFU by two independent scientists. Results were read by a third independent scientist where there was discordance between initial readings.

### S*tatistical analysis*

Linear regression was performed to determine the relationship between N-gene RT-PCR Ct value and either SARS-CoV-2 viral load (reported as RNA copies/mL) or log_10_ TCID_50_/mL. Analytical sensitivity was defined as all samples in a dilution series positive for SARS-CoV-2 antigen, except in the logistic regression analysis, in which each sample was treated as an independent positive (^*^1^*^) or negative (^*^0^*^) result. These regression models were used to determine the 50% antigen test detection rate and 95% confidence intervals (CI) at a given viral load (log_10_ RNA copies/mL). Median value (and corresponding 95% CI) comparing clinical sensitivity and specificity of antigen test positivity to culture infectivity and N-gene RT-PCR Ct values were generated using the Wilson-Brown method. Analysis was performed using GraphPad Prism (v 9.0) and Stata, and data visualisation was performed using GraphPad Prism (v 9.0) and ggplot (v3.3.5) in Rstudio (v1.4.1717) (16).

### Ethics

This study was approved by the Royal Melbourne Hospital Research Ethics Committee (QA2020085).

## RESULTS

### In vitro evaluation of antigen test analytical performance

Overall, 22 rapid antigen kits were assessed for LoD and specificity (Table 1). Nineteen kits consistently detected SARS-CoV-2 antigen at dilutions equivalent to 5.8 × 10^3^ TCID_50_ /mL (1.3 × 10^6^ copies/mL). Six kits consistently detected antigen in samples at a dilution equivalent to 7.2 × 10^2^ TCID_50_ /mL (1.6 × 10^5^ RNA copies/mL), and a further six kits showed a limit of detection at 3.6 × 10^2^ TCID50/mL (8.3 × 10^4^ RNA copies/mL) (Table 1). Linear regression analysis confirmed a significant agreement (R^2^ = 0.9796, P <0.0001) between N-gene Ct and ddPCR-quantified viral load (log_10_ RNA copies/mL) for each sample dilution (Supplementary Figure 2).

**Table 1.** *In vitro* analytical sensitivity of 22 rapid antigen tests to gamma irradiated SARS-CoV-2 Delta variant (B.1.617.2).

|  |  | Analytical sensitivity |  |
| --- | --- | --- | --- |
| Rapid antigen test | Manufacturer | Manufacturer reported<br>(TCID<sub>50</sub>/mL) | VIDRL tested<br>(TCID<sub>50</sub>/mL) |
| Nasal/Nasopharyngeal antigen test |  |  |  |
| OnSite® COVID-19 Ag Point of care test | CTK Biotech Inc | 2.8x10<sup>2</sup> | 1.4x10<sup>3</sup> |
| Ecotest COVID-19 Antigen Nasal Test Kit | Assure Tech (Hangzhou) Co Ltd | 5x10<sup>2</sup> | 1.2x10<sup>4</sup> |
| InnoScreen COVID-19 Antigen Rapid Test Device | Innovation Scientific Pty Ltd | 5x10<sup>2</sup> | 7.2x10<sup>2</sup> |
| SARS-CoV-2 Rapid Antigen Test | SD Biosensor Inc/Roche | 4.9x10<sup>2</sup> | 3.6x10<sup>2</sup> |
| VivaDiag™ SARS-CoV-2 Ag Rapid Test | VivaChek Biotech (Hangzhou) Co Ltd | 6.8x10<sup>2</sup> | 3.6x10<sup>2</sup> |
| Surescreen Diagnostics COVID-19 Antigen Rapid Test Cassette | BTNX Inc | 1x10<sup>3</sup> | 3.6x10<sup>2</sup> |
| NowCheck COVID-19 Antigen Test | BioNote Inc. | 4.9x10<sup>2</sup> | 3.6x10<sup>2</sup> |
| STANDARD™ Q COVID-19 Ag Test | SD Biosensor Inc | 2x10<sup>3</sup> | 7.2x10<sup>2</sup> |
| SARS-CoV-2 Antigen Rapid Test Kit | BIOHIT HealthCare (Hefei) Co Ltd | 0.40x10<sup>2</sup> | 7.2x10<sup>2</sup> |
| BIOCREDIT COVID-19 Ag | RapiGEN Inc | 5.62x10<sup>2</sup> PFU/mL<sup>a</sup> | 2.9x10<sup>3</sup> |
| Wantai SARS-CoV-2 Ag Rapid Test (Colloidal Gold) | Beijing Wantai Biologicalpharmacy Enterprise Co Ltd | 2.3x10<sup>2</sup> | 3.6x10<sup>2</sup> |
| ARISTA™ COVID-19 Antigen Rapid Test | Arista Biotech Pty Ltd | 1x10<sup>2</sup> | 2.9x10<sup>3</sup> |
| BD Veritor™ System for Rapid Detection of SARS-CoV-2 | Becton Dickinson and Company | 1.4x10<sup>2</sup> | 1.2x10<sup>4</sup> |
| GenBody COVID-19 Ag | GenBody Inc | 5.1x10<sup>2</sup> | 1.4x10<sup>3</sup> |
| Novel Coronavirus (SARS-CoV-2) Antigen rapid test | Hangzhou Realy Tech Co Ltd | 1.2x10<sup>3</sup> | 7.2x10<sup>2</sup> |
| Sofia® SARS Antigen FIA | Quidel Corporation | 1.1x10<sup>2</sup> | 2.9x10<sup>3</sup> |
| Abbott PanBio™ COVID-19 Ag Rapid Test Device (Nasal) | Abbott Rapid Diagnostics Jena GmbH | 1.6x10<sup>3</sup> | 7.2x10<sup>2</sup> |
| <sup>a</sup> Testsea SARS-CoV-2 Antigen Test Kit | Hangzhou Testsea Biotechnology Co Ltd | 0.1 ng/mL<sup>a</sup> | 3.6x10<sup>2</sup> |
| CareStart™ COVID-19 Antigen test | AccessBio | 8x10<sup>2</sup> | 1.2x10<sup>4</sup> |
| <sup>a</sup> LYHER Novel Coronavirus (Covid-19) Antigen Test Kit (Colloidal Gold) | Hangzhou Laihe Biotech Co Ltd | 0.5ng/mL<sup>a</sup> | 7.2x10<sup>2</sup> |
| Saliva antigen test |  |  |  |
| 2019-nCoV Ag Saliva Rapid Test Card | Guangzhou Decheng Biotechnology Co Ltd | 1x10<sup>2</sup> | 1.4x10<sup>3</sup> |
| Ecotest COVID-19 Antigen Saliva Test Kit | Assure Tech (Hangzhou) Co Ltd | 0.31x10<sup>2</sup> | 2.9x10<sup>3</sup> |

<sup>a</sup> Values are not directly comparable as reported in other units # Limits set by World Health Organisation (WHO) must be between 10<sup>2</sup> – 10<sup>3</sup> TCID<sub>50</sub>/mL, ideally < 1x10<sup>2</sup>. Results within one log difference of the upper limit were considered ND = Not determined
<sup>a</sup> Values are not directly comparable as reported in other units

Logistic regression analysis demonstrated that eleven kits had a 50% detection probability for viral loads >10^6^ RNA copies/mL, and seven kits had 50% detection probabilities >10^5^ RNA copies/mL, equivalent to approximately 3.7 × 10^3^ TCID_50_/mL (Figure 1 and Supplementary Figure 1). When compared to the manufacturer’s stated sensitivity in the accompanying IFU documentation, 12 kits were less sensitive, although of these, five kits were within a log_10_ difference (Table 1). Three kits provided a limit of detection in ng/mL for recombinant antigen or PFU/mL and therefore could not be compared. None of the kits displayed cross-reactivity with other respiratory viruses (Supplementary Table 4) and no difference in analytical sensitivity was observed between SARS-CoV-2 samples pre- and post-gamma irradiation (Supplementary Table 5).

**Figure 1.**
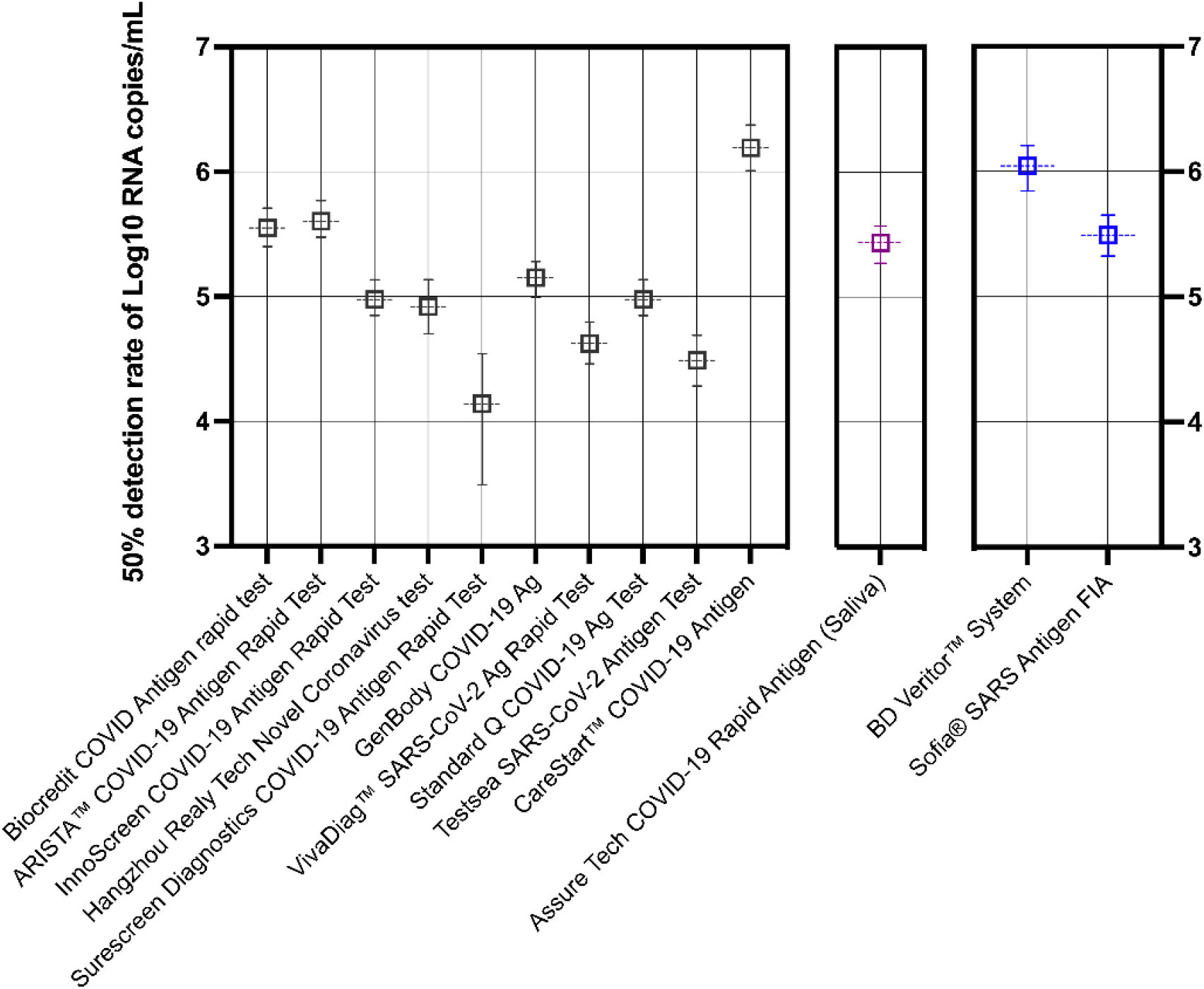
Predictive probability of rapid antigen test positivity against SARS-CoV-2 (B.1.617.2). Thirteen tests were analysed by logistic regression and each result shown is the mean viral load (log_10_ RNA copies/mL) expressed as 50% probability of a positive antigen detection. Note that predictive probabilities could not be determined for some kits, where data showed perfect separation (clear distinction between a positive ^*1*^ and negative ^*1*^ result) and are not depicted above. Error bars represent the 95% confidence interval (CI). Lateral-flow devices (LFDs) are coloured black, saliva-based antigen test highlighted purple and devices requiring a separate device for results interpretation in blue. The Sofia FIA test is a fluorescence-based test.

### Rapid antigen test positivity correlates with culture infectivity

Residual VTM from 78 SARS-CoV-2 RT-PCR-positive clinical specimens were assessed for presence of infectious virus, antigen reactivity (using the Abbott PanBio™ COVID-19 Ag test kit) and quantified for viral RNA copies/mL. The median viral load in the 78 samples was 4.62 × 10^5^ RNA copies/mL (range 1.60 × 10^2^ /mL to 4.41 × 10^8^/mL), with a range of Ct values (median 26.5; range 16.2 to 38.3). In total 41/78 (52.6%; 95% CI 41.6 to 63.3) RT-PCR positive clinical samples were positive using the LFD, of which 40/41 samples (97.6%; 95% CI 86.3 to 100) were also positive for growth in cell culture (Figure 3). Thirty one of the 41 samples with a positive antigen test had a viral load of ≥2 × 10^5^ RNA copies/mL (equivalent N-gene Ct <28). Logistic regression demonstrated a 50% detection probability of cell culture infectivity at a Ct value of 26.6 (95% CI, 24.8 to 28.5) and viral load of 5.9 log_10_ RNA copies/mL (95% CI, 5.3 to 6.5 log_10_ RNA copies/mL) (Figure 2). Viral loads were significantly higher in positive cell cultures and antigen test positive samples compared to culture and antigen test negative samples (Figure 2). The highest Ct value for a positive sample by both antigen test and recoverable infectious virus was 33.4 (Supplementary Table 7).

**Figure 2.**
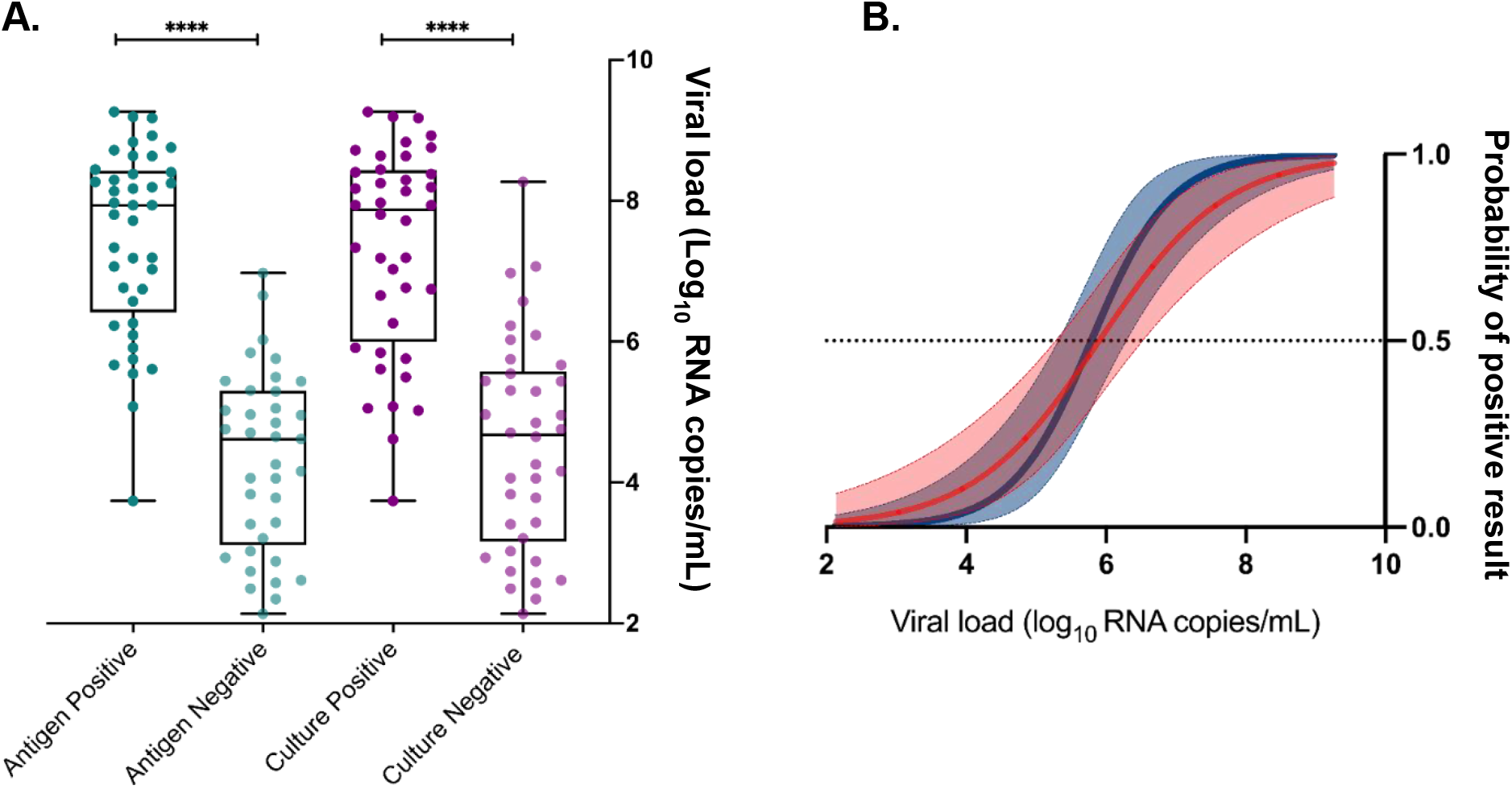
Assessment of viral load, antigen test positivity and cell culture infectivity. Logistic regression models (A) were performed using viral load (log_10_ RNA copies/mL) to calculate the 50% probability of predicting a positive antigen test (dark blue) and positive recovery of infectious virus by cell culture (red). Ninety five percent confidence intervals are shaded and the dotted line represents a 50% positive probability. Virus load in clinical samples (B) was significantly higher for a positive antigen result (dark green) and in samples that recovered infectious virus by cell culture (purple) compared to antigen (light green) and infectivity (light purple) negative samples. ^****^ P value <0.0001.

**Figure 3.**
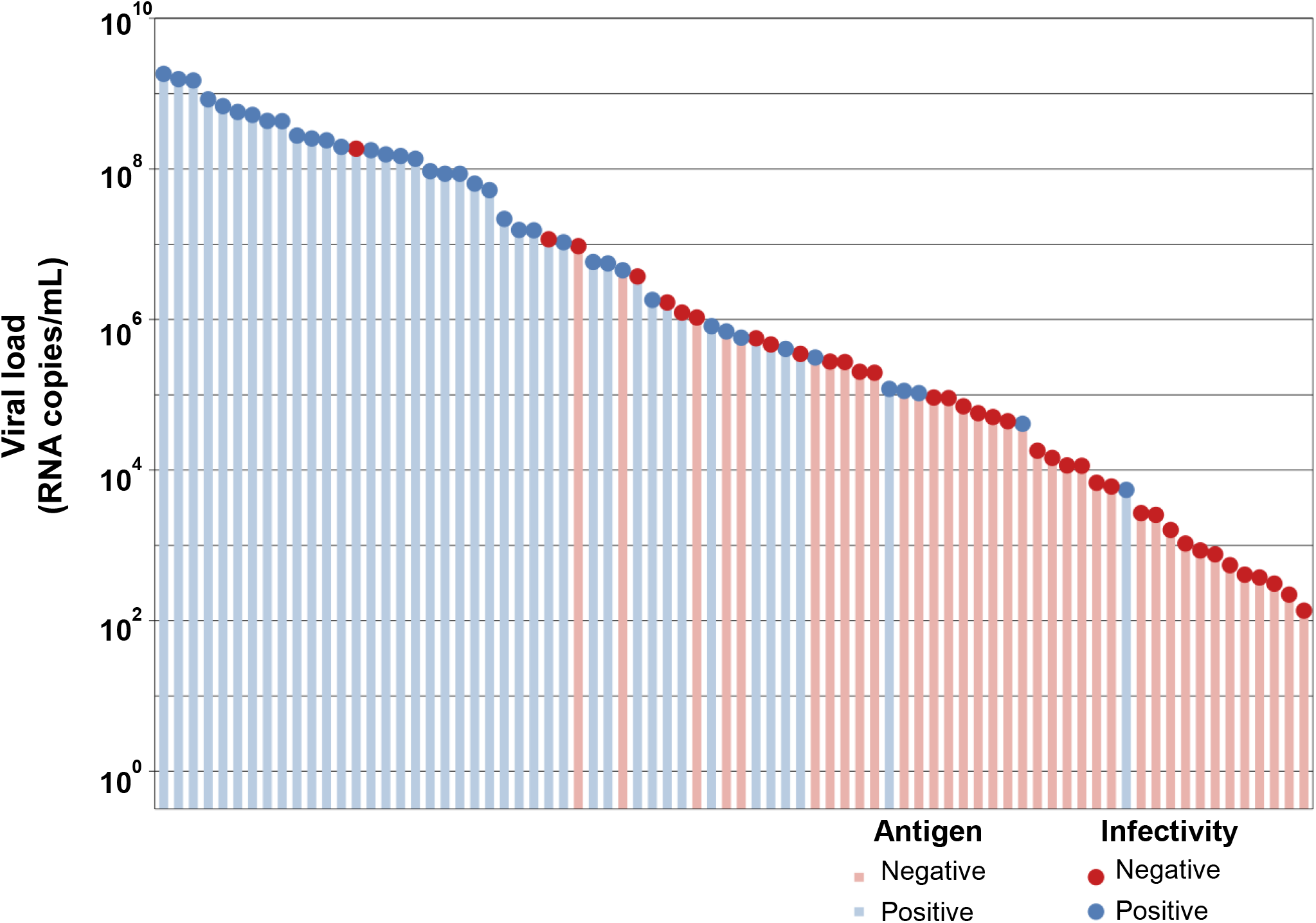
Correlation of antigen test positivity with tissue culture positivity. Clinical naso-oropharyngeal swab samples were submitted for SARS-CoV-2 RT-PCR testing using an in-house N-gene assay. Residual volume was tested for viral infectivity by cell culture (circles), and antigen testing using the Abbott PanBio ™ COVID-19 Ag test kit (bars). Colouring represents a positive sample (blue) or a negative sample (red).

To assess clinical specificity, we tested 100 SARS-CoV-2 negative samples. None of these 100 samples tested positive using the LFD, resulting in 100% specificity compared with RT-PCR (95% CI 96.3% −100%).

## DISCUSSION

Here, we provide information on the comparative analytical performance of twenty-two commercially available LFDs in Australia. As the use of antigen tests increases in Australia, including recent legislative changes allowing home-based antigen testing for COVID-19, knowledge of the performance characteristics is required to help public health agencies, laboratories and the public inform decisions around the selection and implementation of these tests.

All tests were able to detect 1.2 × 10^4^ TCID50/mL, corresponding to approximately 2.6 × 10^6^ viral copies per mL. The World Health Organisation (WHO) suggests a minimum acceptable LoD of 10^2^ −10^3^ TCID50/mL for antigen tests (17), and most (68%) kits, including one saliva antigen test kit, were able to detect 1.4 × 10^3^ TCID_50_/mL, corresponding to a viral load of approximately 3.4 × 10^5^ RNA copies/mL. Our findings are in keeping with other studies that demonstrate comparable sensitivities for a range of commercially available LFDs. For example, Pusken et al. evaluated 31 rapid antigen tests, including 27 LFDs, using a panel of 50 pooled clinical specimens (15). Similar to our findings, these authors demonstrated that the most sensitive kits were able to detect approximately 7.5 × 10^4^ RNA copies/mL, and the least sensitive kit approximately 2.3 × 10^7^ RNA copies/mL (15). In our study, detection limits ranged from approximately 8.3 × 10^4^ (six kits) to 2.6 × 10^6^ RNA copies/mL (all 22 kits). Further, Corman et al. evaluated seven commercially available LFDs and demonstrated that six kits were able to detect between 2.07 × 10^7^ and 2.86 × 10^6^ RNA copies/mL (18). Importantly however, studies reporting analytical sensitivity of LFDs vary across several parameters, including sample matrix (e.g. VTM or saline) (19), antigenic source (e.g. recombinant antigen; virus inactivation method; swab type) (20); reporting format (e.g. PFU/mL; TCID_50_; copies/mL) and different SARS-CoV-2 variants (19, 20). Here, we correlate antigen test positivity with: (i) Ct values; (ii) RNA copies/mL; and (iii) TCID_50_/mL to provide a systematic comparison of quantitative laboratory parameters. Our finding of differential analytical sensitivity of antigen kits using standardised panels reiterates the need for ongoing post-market validation data, including regular assessment of batch variation, to help guide the deployment of these tests.

Similar to other studies (21), we found high analytical specificity of antigen tests, with no cross-reactivity for any antigen tests against a panel of other respiratory viruses. Specificity was also high (100%) for the Abbott PanBio™ COVID-19 Ag test kit against clinical samples, in keeping with other findings, and with WHO recommendations of a minimum specificity of ≥97% for rapid antigen tests (7, 17). Importantly however, even with high specificity, the positive predictive value of antigen testing will be low when there is minimal community transmission; in these circumstances, confirmatory RT-PCR testing is strongly recommended (22).

Previous work has demonstrated a correlation between respiratory tract specimens with higher SARS-CoV-2 viral loads and infectivity in cell culture (8, 23, 24, 25). For example, Pickering et al. found that approximately 95% of clinical samples that yielded a positive viral culture had a Ct value of less than 25 (8). Further, when culture positivity was used as the reference standard, rather than RT-PCR, the sensitivity of antigen testing increased to at 94.7% (8). Similarly, Pekosz et al., found that antigen testing (using the BD Veritor system) demonstrated a higher positive percent agreement (PPA) (∼90%) than RT-PCR (∼70%) when using positive viral culture as the reference gold standard (26). In our study, 40/41 (98%) of samples with a positive viral culture also had a positive antigen test, with 95% of these samples having a Ct of less than 30. However, we also observed 8/40 (20%) samples that had a positive viral culture with Ct values of ≥ 27; this may reflect differences in RT-PCR assays and in cell lines used for viral culture across different studies. In our work we utilised an in-house RT-PCR assay and Calu-3 cells for viral culture, whereas Pickering et al. used the AusDiagnostics multiplexed-tandem PCR assay and Vero E6 cells, and Pekosz et al. used the Quidel Lyra SARS-CoV-2 Assay and VeroE6TMPRSS2 cells (8, 26). Further, in our study, the majority (65.5%) of clinical samples consisted of the SARS-CoV-2 Delta variant, the predominant global variant, whereas other similar studies have focused on previously circulating variants, such as B.1.1.7 (Alpha variant) (8, 27). These study differences highlight the need for standardised approaches and protocols for studies assessing viral infectivity, particularly if results are subsequently translated into public health or infection control policies.

A limitation of our study, and indeed a limitation of several other laboratory studies assessing antigen test kits, include the use of spiked virus in VTM for kit evaluation, rather than the use of swabs directly placed in manufacturer-provided buffer. However, to try and maximise yield from samples, we tested samples without a freeze-thaw step, as freezing has been shown to reduce yield of infectious virus (18, 28). Although we assessed the analytical characteristics of over twenty assays, a further limitation is that we only evaluated the clinical sensitivity of one antigen test, namely the Abbott PanBio™ COVID-19 Ag test assay. However, recent work has highlighted the widespread use of this kit, with 39 published datasets utilising the Abbott PanBio™ COVID-19 Ag test assay (21), meaning that our findings will have broad applicability and relevance.

In summary, our data describe the performance characteristics of 22 antigen test kits against the SARS-CoV-2 Delta variant using a standardised evaluation panel. We demonstrate marked variability between test kits, and variability between reported and observed sensitivities, although most (86.4%) were able to meet WHO recommended minimum standards for detection. In addition, we further corroborate the hypothesis that antigen test positivity generally correlates with positive viral culture and by extension with presence of infectious viral particles) and that a positive antigen test result could be used as an adjunct to determine SARS-CoV-2 transmission risk. Collectively, our data provide valuable information to help guide antigen test selection and deployment and highlight the need for ongoing systematic post-market evaluation of antigen test kits, ideally using standardised reagents and protocols.

## Data Availability

All data produced in the present study are available upon reasonable request to the authors

## Acknowledgements

This work was supported by the Department of Health Victoria, Australia. We thank the study participants for their contributions.

## Notes

### Competing Interest Statement

The authors have declared no competing interest.

### Funding Statement

This study was funded by the Department of Health and Human Services Victoria, Australia

### Author Declarations

The Human Research Ethics Committee of the Royal Melbourne Hospital gave ethical approval for this work

