## Supplemental tables and figures for "Assessment of twenty-two SARS-CoV-2 rapid antigen tests against SARS-CoV-2: A laboratory evaluation study"

**SUPPLEMENTARY APPENDIX**

#### SUPPLEMENTARY METHODS

##### *SARS-CoV-2 Cell culture*

SARS-CoV-2 (VIC/18440/2021) was originally isolated from residual VTM from a RT-PCR-positive swab collected on the 17th April, 2021 at VIDRL. Whole genome sequencing and bioinformatic analysis was performed by the Microbiological Diagnostic Unit Public Health Laboratory using previously described methods (29, 30) and determined the lineage as B.1.617.2 (Delta) (GISAID Accession ID: EPI_ISL_1913206). To prepare the stock used in this study, 100µl of virus isolate was added to 2 x T25 flasks of Calu-3 cells grown to 80% confluency and allowed to absorb for 30 min at 37°C. 10mL of Calu-3 infection media (1X MEM *Life Technologies*, 1% Penicillin/Streptomycin *Sigma-Aldrich*, 1X GlutaMax *Life Technologies,* and 15mM HEPES *Sigma-Aldrich,* 0.8% gentamicin, 4µg/mL Trypsin) was added to each flask and cells incubated at 37°C. When CPE was observed (approximately 72 hours post infection) supernatant from each flask was harvested, combined and cell debris removed by centrifugation. Aliquots of virus were stored at -80°C.

For the clinical culture infectivity assay, SARS-CoV-2 RT-PCR positive samples were filtered through a 0.45µm filter to prevent microbial growth during culture. 150µL of filtered VTM was absorbed onto a confluent monolayer of human airway epithelial cells (Calu-3) in a single well of a 24-well plate for 30 minutes. Following inoculation, 500µL of Calu-3 infection media (DMEM/F-12, supplemented with GlutaMAX *Gibco*, 2% heat inactivated FBS) was added to each well and cells incubated at 37°C for ~96 hours. The presence of virus-induced cytopathic effect (CPE) was recorded for each sample and RT-PCR used to confirm culture infectivity status for each sample. All SARS-CoV-2 culture was performed in a physical containment level 3 (PC3) laboratory.

***Quantitation of SARS-CoV-2 by $TCID_{50}$ assay***

Quantification of SARS-CoV-2 stock was by $TCID_{50}$ assay. In a PC3 laboratory, 10-fold dilutions ($10^{-4}$-$10^{-8}$) of SARS-CoV-2 (VIC/18440/2021, described above) were prepared with infection media (DMEM/F-12, supplemented with GlutaMAX *Gibco*, 2% heat inactivated FBS). Duplicate assays were prepared with 150µL of each dilution added either as 4 or 6 replicates in a 24-well plate containing Calu-3 cells grown to ~90% confluency. Virus was allowed to absorb for 30 minutes at 37°C before an additional 350µl of infection media was added and plates returned to incubate at 37°C. Approximately 96 hours post infection, cytopathic effect was observed. The $TCID_{50}$ value for the stock virus was obtained by averaging the replicate results calculated using the Spearman & Karber algorithm (31).

***Nucleic acid extraction, reverse transcription, and in-house SARS-CoV-2 N-gene RT-PCR***

Nucleic acid was extracted using a QIAamp 96 virus QIAcube HT kit (Qiagen, Germany) on the QIAcube HT System (Qiagen, Germany) according to manufacturer's instructions. In brief, a 200µL volume of each sample was used for extraction and eluted in a 60µL volume of elution buffer into a 96-well elution tray. Reverse transcription of viral RNA to complimentary DNA (cDNA) was performed using Bioline SensiFAST cDNA synthesis kit (Bioline Reagents Ltd. UK) according to manufacturer's instruction. In brief, a 10µL volume of eluted nucleic acid was added to 10µL of SensiFAST cDNA reaction mix, heat linearized at 25˚C for 10 minutes then incubated at 42˚C for 15 minutes before denaturation at 85˚C for 5 minutes on an Applied Biosystems SimpliAmp thermal cycler (Thermo Fisher Scientific, Singapore).

An in-house real-time RT-PCR assay targeting the SARS-CoV-2 N-gene was used. Primers and probe sequences targeting the N-gene have been previously described (32). Briefly, 3µl of cDNA was added to a commercial real-time PCR master mix (PrecisionFast qPCR Master Mix with LowRox, Primer Design Ltd.,UK) in a 20µl reaction mix containing primers and probe with a final concentration of 0.9µM and 0.2µM for primer and the probe, respectively. An in-house positive extraction control, a negative control and a positive control were included with each PCR run. Thermal cycling and real-time PCR analysis for all assays were performed on an ABI 7500 FAST real-time PCR system (Applied Biosystems, Foster City, CA) with the thermal cycling profile: 95°C for 2 min, followed by 45 PCR cycles of 95°C for 5 sec and 60°C for 25 sec. A positive result was interpreted from uniform S-shaped amplification profiles where the threshold line was set to intersect with the mid-point of the linear amplification phase of the amplification-curve. A result less than or equal to a cycle threshold (Ct) value of 40 was considered positive.

***Preparation and Droplet Digital PCR quantitation of SARS-CoV-2 standard curve***

A 12-point 2-fold dilution series (1/800 to 1/1,638,400 dilution) of gamma irradiated SARS-CoV-2 (VIC/18440/2021) stock was prepared in VTM (1X MEM *Gibco* supplemented with 2% heat inactivated FBS, 1X GlutaMax, 15mM HEPES, 0.1mg/mL penicillin/streptomycin, 2.5µg/mL amphotericin B and 20µg/mL neomycin). For the first and final three dilution points, six replicates, from three separate nucleic acid extraction runs, was performed using the QIAamp 96 virus QIAcube HT kit on the QIAcube HT system. For all other dilution points, twelve replicates from three separate nucleic extraction runs were used. The purified nucleic acid was then converted to cDNA as described above. Prior to performing droplet digital PCR (ddPCR) the cDNA samples were first tested in an in-house qualitative real-time PCR assay targeting the N-gene described above.

For ddPCR, primers and probe from the qualitative N-gene real-time RT-PCR was used to prepare a ddPCR Supermix (ddPCR Supermix for Probes, Bio-Rad, Bio-Rad Laboratories. Inc, USA). The Supermix was prepared with the same concentration of primers and probe as the qualitative assay. 3µl of cDNA template was added to the Supermix for a final reaction volume of 25µl. Droplets were generated from 20µl of this mixture on the QX200 Droplet Generator (Bio-Rad, USA) as per manufacturer's instructions and run on a thermal cycler (Thermo Scientific Arktik Thermal Cycler, Thermo Fisher Scientific) with thermal cycling conditions: 95˚C for 10 min, followed by 40 cycles of 95˚C for 30 sec and 60˚C for 1 min, 98˚C for 10 min and hold 4˚C. A QX200 Droplet Reader (Bio-Rad, USA) was used to take fluorescence readings. Each replicate per dilution point was tested in the ddPCR, RNA copies/mL calculated for each point and a quantitative linear regression standard curve established. Five nucleic acid extracted replicates of a commercially available SARS-CoV-2 reference material (AccuPlex SARS-CoV-2 Reference Material Kit, SeraCare Life Sciences, Inc. Milford, MA USA) was used to verify the performance of the standard curve.

***Preparation of specificity panel***

The panel of 'distractor' viruses was prepared using viral isolates stored at VIDRL. Specifically, viruses and cell lines were: Influenza A H1N1 and Influenza B (MDCK cells), Respiratory Syncytial Virus (RSV) (Hep-2 cells), Adenovirus 3 (A549 cells), Rhinovirus (H1HeLa cells), human coronaviruses (OC43 and 229E), Herpes Simplex Virus 1 (HSV-1) and Cytomegalovirus (CMV) (all grown in Human Embryonic Lung cells), and Parainfluenza virus

3 (MK2 cells). Virus stocks were diluted 1:100 in VTM or pooled saliva to achieve a Ct ≤25. For HSV-1, samples were diluted 1:10 in VTM or pooled saliva. Viral RNA was purified and reverse transcribed as described above. A list of primer and probe sequences for each virus is provided (Supplementary Table 3). Each virus in the panel was quantified using ddPCR. Primers and probe from the qualitative real-time RT-PCR were used to prepare a ddPCR Supermix (ddPCR Supermix for Probes, Bio-Rad, Bio-Rad Laboratories. Inc, USA). The Supermix was prepared with the same concentration of primers and probe as the qualitative assay. Three microliters of cDNA template was added to 22µl of the Supermix for a final reaction volume of 25µl. For DNA viruses 4µl of extracted DNA was used directly. Droplets were generated from 20µl of this mixture on the QX200 Droplet Generator (Bio-Rad, USA) as per manufacturer's instructions and run on a thermal cycler (Thermo Scientific Arktik Thermal Cycler, Thermo Fisher Scientific) with thermal cycling conditions: 95˚C for 10 min, followed by 40 cycles of 95˚C for 30 sec and 60˚C for 1 min, 98˚C for 10 min and hold 4˚C. A QX200 Droplet Reader (Bio-Rad, USA) was used to take fluorescence readings. RNA copies/mL were calculated and reported for each virus (Supplementary Table 4).

***Quantitation of SARS-CoV-2-positive clinical samples***

For clinical samples, viral loads of eluted swabs were quantified to allow correlation with cell culture infectivity and antigen test reactivity. Briefly, extracted nucleic acid was reverse transcribed as described above. An in-house RT-PCR assay detecting the nucleocapsid gene of SARS-CoV-2 (described above) was performed on each sample and RNA copy number/mL was quantified using the following linear regression formula: $y = -0.2907x + 13.349$. For each SARS-CoV-2-positive clinical sample, the Ct value and RNA copies/mL values were recorded.

**Supplementary Table 1. Antigen tests evaluated in this study, with assay-specific sample and diluent volumes.**

| Test | Manufacturer | Time (mins) | Buffer volume (μL) | | |
|---|---|---|---|---|---|
| | | | Extraction | Sample | Assay input |
| **Nasal/Nasopharyngeal antigen test** | | | | | |
| OnSite® COVID-19 Ag Point of care test / Aria® COVID-19 Ag Rapid Test | CTK Biotech Inc (USA) | 15-20 | NS | 90 | 90 |
| Ecotest COVID-19 Antigen Nasal Test Kit | Assure Tech (Hangzhou) Co Ltd (China) | 15 | 150 | 150 | 80 |
| InnoScreen COVID-19 Antigen Rapid Test Device | Innovation Scientific Pty Ltd (Australia) | 15 | 400 | 400 | 100 |
| SARS-CoV-2 Rapid Antigen Test | SD Biosensor Inc (Korea - Republic of) | 15-30 | 300 | 350 | 50 |
| VivaDiag™ SARS-CoV-2 Ag Rapid Test | VivaChek Biotech (Hangzhou) Co Ltd (China) | 15-20 | 300 | 300 | 60 |
| Surescreen Diagnostics COVID-19 Antigen Rapid Test Cassette | BTNX Inc (Canada) | 15 | 300 | 300 | 80 |
| NowCheck COVID-19 Antigen Test | BioNote Inc (Korea - Republic of) | 15-30 | 300 | 300 | 50 |
| STANDARD™ Q COVID-19 Ag Test | SD Biosensor Inc (Korea - Republic of) | 15-20 | 300 | 300 | 50 |
| SARS-CoV-2 Antigen Rapid Test Kit * # | BIOHIT HealthCare (Hefei) Co Ltd (China) | 15 | 300 | 300 | 75 |
| BIOCREDIT COVID-19 Ag | RapiGEN Inc (Korea - Republic of) | 10-15 | 400 | 400 | 100 |
| Wantai SARS-CoV-2 Ag Rapid Test (Colloidal Gold) | Beijing Wantai Biologicalpharmacy Enterprise Co Ltd (China) | 20-30 | 60 | 300 | 100 |
| ARISTA™ COVID-19 Antigen Rapid Test | Arista Biotech Pte Ltd (Singapore) | 15-30 | 500 | 500 | 50 |
| BD Veritor™ System for Rapid Detection of SARS-CoV-2 * | Becton Dickinson and Company (United States Of America) | 15 | 325 | 325 | 70 |
| GenBody COVID-19 Ag | GenBody Inc (Korea - Republic of) | 15-20 | 400 | 400 | 100 |
| Novel Coronavirus (SARS-CoV-2) Antigen rapid test | Hangzhou Realy Tech Co Ltd (China) | 10-20 | 250 | 250 | 90 |
| Sofia® SARS Antigen FIA * # | Quidel Corporation (United States Of America) | 15 | NS | 250 | 120 |
| Panbio™ COVID-19 Ag Rapid Test Device (Nasal) | Abbott Rapid Diagnostics Jena GmbH (Germany) | 15 | 150 | 150 | 130 |
| Testsea SARS-CoV-2 Antigen Test Kit | Hangzhou Testsea Biotechnology Co Ltd (China) | 10-15 | 300 | 300 | 55 |
| CareStart™ COVID-19 Antigen test | AccessBio | 10 | 450 | 450 | 70 |
| LYHER Novel Coronavirus (Covid-19) Antigen Test Kit (Colloidal Gold) | Hangzhou Laihe Biotech Co Ltd (China) | 15 | 250 | 250 | 100 |
| **Saliva antigen test** | | | | | |
| 2019-nCoV Ag Saliva Rapid Test Card | Guangzhou Decheng Biotechnology Co Ltd (China) | 10 | 0 | 800 | N/A |
| Ecotest COVID-19 Antigen Saliva Test Kit | Assure Tech (Hangzhou) Co Ltd (China) | 15 | 0 | 400 | N/A |

* Requires an independent reading device to interpret results

### Fluorescent test

N/A – Not applicable. Test was incubated inverted in the sample tube for 2 minutes.

N/S - Not specified

**Supplementary Table 2. Analytical sensitivity panel.** Two-fold dilutions of gamma-irradiated SARS-CoV-2 for limit of detection analysis expressed as cell culture infectivity ($TCID_{50}$) and viral load (RNA copies/mL), calculated from a standard curve using N-gene Ct values. Dilutions 1:800 – 1:204,800 were prepared in quadruplicate, with the remaining dilutions tested in duplicate.

| **Dilution** | **Ct (N-gene)** | **$TCID_{50}$/mL** | **Viral load (RNA copies/mL)** |
|---|---|---|---|
| 100 | 20.6 | $9.3x10^4$ | $2.2x10^7$ |
| 200 | 21.7 | $4.6x10^4$ | $1.1x10^7$ |
| 400 | 22.7 | $2.3x10^4$ | $5.5x10^6$ |
| 800 | 23.9 | $12x10^4$ | $2.6x10^6$ |
| 1,600 | 24.8 | $5.8x10^3$ | $1.3x10^6$ |
| 3,200 | 26.3 | $2.9x10^3$ | $5.2x10^5$ |
| 6,400 | 26.9 | $1.4x10^3$ | $3.4x10^5$ |
| 12,800 | 28.0 | $7.2x10^2$ | $1.6x10^5$ |
| 25,600 | 29.0 | $3.6x10^2$ | $8.3x10^4$ |
| 51,200 | 30.0 | $1.8x10^2$ | $4.1x10^4$ |
| 102,400 | 30.9 | $0.90x10^2$ | $2.3x10^4$ |
| 204,800 | 31.9 | $0.45x10^2$ | $1.2x10^4$ |
| 409,600 | 33.1 | $0.23x10^2$ | $5.2x10^3$ |
| 819,200 | 34.2 | $0.11x10^2$ | $2.6x10^3$ |
| 1,638,400 | 35.2 | $0.06x10^2$ | $1.3x10^3$ |
| Negative control | ND | 0 | ND |

$TCID_{50}$ – Median tissue culture infectious dose
ND – Not detected

**Supplementary Table 3. Primers used for quantitation of distractor viruses by droplet digital RT-PCR.**

| Distractor | Forward Primer | Reverse Primer | Probe |
|---|---|---|---|
| Influenza A | MGAGGTCGAAACGTAYGTTCTCT | GTCTTGTCTTTAGCCAYTCCATGA | CCCCCTCAAAGCCGA |
| Influenza B | ATGGATACAAGTCCTTATCAACTCTGC | TTCATTAARACGCTCGAAGAG | CCA**T**CT**T**CT**T**CA**T**CCTCCACT |
| Respiratory Syncytial Virus | GAGTTGAAGGRATYTTTGCAGGA<br>GAGTTGAAGGRATYTTTGCAGGAT | AAAACTCCCCATCTTAGCATTACTTG<br>CCCCCACCGTAACATCACTT | TTATGAATGCCTATGGTKCAG |
| Rhinovirus | AGCCTGCGTGGCTGCCTG<br>CCTGCGTGGCGGCCARC | CCCAAGTAGTYGGTCCCRTCC | TCCTCCGGCYCCTGAATG |
| HCoV-229E | TCACATGTTGTACGGCTAGTGATAAA | ACCCACCATTTGAATAAACAACCT | AGCAAGCTCATTACTAAGTCTA |
| HCoV-OC43 | AAATTTTATGGTGGCTGGAATAATATGTT | TAGGCATAGCTCTRTCACAYTT<br>TTGGCATRGCACGATCACAYTT | TGGGTTGGGATTATC |
| Adenovirus 3 | TGGKCDTACATGCACATCKC | GGGTTTCTAAACTTGTTATTCAGG<br>GGGTTTCTAAACTTGTTSCCCAKR | CGC**C**TC**G**GA**G**TA**C**CTG<br>TGC**T**TC**G**GA**G**TA**T**CTG |
| Human Simplex Virus 1 | ACCACGAGACCGACATGGAG | GTTGTACTTGAGGTCGGTGGTG | CCAAYGCCGCGACCCGCACG |
| Cytomegalovirus | CCGGCAAGCTCTTTATGCA | TGGGACACAACACCGTAAAGC | ACG**A**TG**A**CC**C**GC**A**ACC |
| Parainfluenza 3 | GATTAGAGGCTTTCAGACAAGATGG | CTCTGTTGAGACCGCATGATTG | CCAATCTGATCCACTGTGTCACCGCTCA |
| SARS-CoV-2 (N-gene) | CACATTGGCACCCGCAATC | GAGGAACGAGAAGAGGCTTG | ACTTCCTCAAGGAACAACATTGCCA |

Probes including locked nucleic acid bases (LNAs) have the LNA indicated with bold font

**Supplementary Table 4. Specificity evaluation of rapid antigen tests.** A panel of viruses were tested against each antigen test to evaluate cross-reactivity. Each sample is expressed in viral load (copies/mL) and assessed in duplicate.

| | | | VTM PANEL | | | | | | | | | | | | | | | | | | | | SALIVA PANEL | |
|---|---|---|---|---|---|---|---|---|---|---|---|---|---|---|---|---|---|---|---|---|---|---|---|---|
| **Distractor virus** | **Ct** | **Viral load (copies/mL)** | Assure Tech Ecotest COVID-19 Rapid Ag Test (Naso) | BD Veritor™ System for Rapid Detection of SARS-CoV-2 | Biocredit COVID Ag rapid test | ARISTA™ COVID-19 Ag Rapid Test | InnoScreen COVID-19 Ag Rapid Test Device | OnSite® COVID-19 Ag Point of care test | Hangzhou Realy Tech Novel Coronavirus (SARS-CoV-2) Ag rapid test | Wantai SARS-CoV-2 Ag Rapid Test (Colloidal Gold) | Sofia® SARS Ag FIA | Surescreen Diagnostics COVID-19 Ag Rapid Test Cassette | LYHER Novel Coronavirus (Covid-19) Ag Test Kit (Colloidal Gold) | GenBody COVID-19 Ag | VivaDiag™ SARS-CoV-2 Ag Rapid Test | SD Biosensor Inc/Roche SARS-CoV-2 Rapid Ag Test (NP) | STANDARD Q COVID-19 Ag Tests | BIOHIT HealthCareSARS-CoV-2 Ag Rapid Test | NowCheck COVID-19 Ag Test | CareStart™ COVID-19 Antigen test | Panbio COVID-19 Rapid Test Device (nasal) | Testsea SARS-CoV-2 Ag Test Kit | Assure Tech Ecotest COVID-19 Rapid Ag SALIVA | 2ND19-nCoV Ag Saliva Rapid Test Card |
| Influenza A | 25 | 1,343,063 | ND<br>ND | ND<br>ND | ND<br>ND | ND<br>ND | ND<br>ND | ND<br>ND | ND<br>ND | ND<br>ND | ND<br>ND | ND<br>ND | ND<br>ND | ND<br>ND | ND<br>ND | ND<br>ND | ND<br>ND | ND<br>ND | ND<br>ND | ND<br>ND | ND<br>ND | ND<br>ND | ND<br>ND | ND<br>ND |
| Influenza B | 24 | 3,551,813 | ND<br>ND | ND<br>ND | ND<br>ND | ND<br>ND | ND<br>ND | ND<br>ND | ND<br>ND | ND<br>ND | ND<br>ND | ND<br>ND | ND<br>ND | ND<br>ND | ND<br>ND | ND<br>ND | ND<br>ND | ND<br>ND | ND<br>ND | ND<br>ND | ND<br>ND | ND<br>ND | ND<br>ND | ND<br>ND |
| RSV | 25 | 1,617,375 | ND<br>ND | ND<br>ND | ND<br>ND | ND<br>ND | ND<br>ND | ND<br>ND | ND<br>ND | ND<br>ND | ND<br>ND | ND<br>ND | ND<br>ND | ND<br>ND | ND<br>ND | ND<br>ND | ND<br>ND | ND<br>ND | ND<br>ND | ND<br>ND | ND<br>ND | ND<br>ND | ND<br>ND | ND<br>ND |
| Rhinovirus | 23 | 5,486,250 | ND<br>ND | ND<br>ND | ND<br>ND | ND<br>ND | ND<br>ND | ND<br>ND | ND<br>ND | ND<br>ND | ND<br>ND | ND<br>ND | ND<br>ND | ND<br>ND | ND<br>ND | ND<br>ND | ND<br>ND | ND<br>ND | ND<br>ND | ND<br>ND | ND<br>ND | ND<br>ND | ND<br>ND | ND<br>ND |
| Cov-229E | 25 | 3,626,625 | ND<br>ND | ND<br>ND | ND<br>ND | ND<br>ND | ND<br>ND | ND<br>ND | ND<br>ND | ND<br>ND | ND<br>ND | ND<br>ND | ND<br>ND | ND<br>ND | ND<br>ND | ND<br>ND | ND<br>ND | ND<br>ND | ND<br>ND | ND<br>ND | ND<br>ND | ND<br>ND | ND<br>ND | ND<br>ND |
| Cov-OC43 | 19 | 38,190,000 | ND<br>ND | ND<br>ND | ND<br>ND | ND<br>ND | ND<br>ND | ND<br>ND | ND<br>ND | ND<br>ND | ND<br>ND | ND<br>ND | ND<br>ND | ND<br>ND | ND<br>ND | ND<br>ND | ND<br>ND | ND<br>ND | ND<br>ND | ND<br>ND | ND<br>ND | ND<br>ND | ND<br>ND | ND<br>ND |
| Adenovirus | 19 | 85,065,000 | ND<br>ND | ND<br>ND | ND<br>ND | ND<br>ND | ND<br>ND | ND<br>ND | ND<br>ND | ND<br>ND | ND<br>ND | ND<br>ND | ND<br>ND | ND<br>ND | ND<br>ND | ND<br>ND | ND<br>ND | ND<br>ND | ND<br>ND | ND<br>ND | ND<br>ND | ND<br>ND | ND<br>ND | ND<br>ND |
| Human Simplex Virus 1 | 23 | 4,066,000 | ND<br>ND | ND<br>ND | ND<br>ND | ND<br>ND | ND<br>ND | ND<br>ND | ND<br>ND | ND<br>ND | ND<br>ND | ND<br>ND | ND<br>ND | ND<br>ND | ND<br>ND | ND<br>ND | ND<br>ND | ND<br>ND | ND<br>ND | ND<br>ND | ND<br>ND | ND<br>ND | ND<br>ND | ND<br>ND |
| Cytomegalovirus | 24 | 486,850 | ND<br>ND | ND<br>ND | ND<br>ND | ND<br>ND | ND<br>ND | ND<br>ND | ND<br>ND | ND<br>ND | ND<br>ND | ND<br>ND | ND<br>ND | ND<br>ND | ND<br>ND | ND<br>ND | ND<br>ND | ND<br>ND | ND<br>ND | ND<br>ND | ND<br>ND | ND<br>ND | ND<br>ND | ND<br>ND |
| Parainfluenza 3 virus | 23 | 22,051,875 | ND<br>ND | ND<br>ND | ND<br>ND | ND<br>ND | ND<br>ND | ND<br>ND | ND<br>ND | ND<br>ND | ND<br>ND | ND<br>ND | ND<br>ND | ND<br>ND | ND<br>ND | ND<br>ND | ND<br>ND | ND<br>ND | ND<br>ND | ND<br>ND | ND<br>ND | ND<br>ND | ND<br>ND | ND<br>ND |

**Supplementary Table 5. Assessment of antigen test positivity of pre- and post-gamma irradiation of SARS-CoV-2.** Ten-fold serial dilutions of SARS-CoV-2 were prepared from stock material before and after exposure to gamma-irradiation and tested in quadruplicate using the Abbott Panbio antigen Rapid test device.

| Dilution | Pre-Gamma irradiation | | | | Post-Gamma irradiation | | | |
|---|---|---|---|---|---|---|---|---|
| 100 | + | + | + | + | + | + | + | + |
| 1000 | + | + | + | + | + | + | + | + |
| 10000 | + | + | + | + | + | + | + | + |
| 100000 | - | - | - | - | + | - | - | - |
| 1000000 | - | - | - | - | - | - | - | - |

#### Supplementary Table 6. Binomial logistic regression analysis for antigen test sensitivity

| Sample | Ct (N) | Viral Load (RNA copies/mL) | Antigen test | Infectivity result | Lineage |
|---|---|---|---|---|---|
| **1** | 23.3 | 3.79E+06 | + | + | N/A |
| **2** | 27.6 | 2.10E+05 | - | - | B.1.617.2 |
| **3** | 19.3 | 5.62E+07 | + | + | B.1.617.2 |
| **4** | 27.0 | 3.07E+05 | + | + | B.1.617.2 |
| **5** | 20.7 | 2.17E+07 | + | + | B.1.1.7 |
| **6** | 20.6 | 2.34E+07 | + | + | B.1.1.7 |
| **7** | 26.9 | 3.50E+05 | + | - | B.1.1.7 |
| **8** | 24.9 | 1.27E+06 | + | + | B.1.1.7 |
| **9** | 16.3 | 4.16E+08 | + | + | B.1.617.2 |
| **10** | 34.1 | 2.68E+03 | - | - | B.1.617.2 |
| **11** | 23.6 | 3.11E+06 | - | + | B.1.617.2 |
| **12** | 35.4 | 1.11E+03 | - | - | B.1.617.2 |
| **13** | 23.6 | 3.11E+06 | + | + | N/A |
| **14** | 26.3 | 5.09E+05 | - | + | B.1.617.2 |
| **15** | 19.2 | 5.92E+07 | + | + | B.1.617.2 |
| **16** | 29.1 | 7.85E+04 | - | - | B.1.617.2 |
| **17** | 20.7 | 2.18E+07 | + | + | B.1.617.2 |
| **18** | 29.1 | 7.63E+04 | - | - | B.1.617.2 |
| **19** | 32.4 | 8.32E+03 | - | - | B.1.617.2 |
| **20** | 20.4 | 2.66E+07 | + | + | B.1.617.2 |
| **21** | 25.5 | 8.36E+05 | + | - | B.1.617.2 |
| **22** | 37.1 | 3.55E+02 | - | - | N/A |
| **23** | 32.7 | 6.92E+03 | - | - | B.1.617.2 |
| **24** | 37.4 | 3.03E+02 | - | - | N/A |
| **25** | 30.6 | 2.89E+04 | - | - | B.1.617.2 |
| **26** | 36.9 | 4.26E+02 | - | - | N/A |
| **27** | 38.1 | 1.88E+02 | - | - | N/A |
| **28** | 37.3 | 3.28E+02 | - | - | N/A |
| **29** | 28.1 | 1.49E+05 | + | - | N/A |
| **30** | 29.9 | 4.45E+04 | - | - | B.1.617.2 |
| **31** | 33.6 | 3.78E+03 | - | - | B.1.617.2 |
| **32** | 36.6 | 5.12E+02 | - | - | N/A |
| **33** | 19.2 | 5.94E+07 | + | + | B.1.617.2 |
| **34** | 33.6 | 3.81E+03 | - | - | B.1.617.2 |
| **35** | 30.9 | 2.35E+04 | - | - | B.1.617.2 |
| **36** | 32.8 | 6.36E+03 | - | - | B.1.617.2 |
| **37** | 38.1 | 1.94E+02 | - | - | B.1 |
| **38** | 28.1 | 1.54E+05 | - | + | AY.4 |
| **39** | 35.7 | 9.44E+02 | - | - | AY.12 |
| **40** | 27.1 | 3.01E+05 | - | + | B.1.617.2 |
| **41** | 38.3 | 1.60E+02 | - | - | N/A |
| **42** | 32.4 | 8.73E+03 | - | - | N/A |
| **43** | 17.8 | 1.54E+08 | + | + | B.1.617.2 |
| **44** | 19.7 | 4.22E+07 | + | + | B.1.617.2 |
| **45** | 29.9 | 4.52E+04 | - | - | B.1.617.2 |
| **46** | 30.3 | 3.36E+04 | - | - | N/A |
| **47** | 24.3 | 1.94E+06 | - | + | B.1.617.2 |
| **48** | 22.6 | 5.85E+06 | + | + | B.1.617.2 |
| **49** | 27.1 | 2.96E+05 | + | - | AY.4 |
| **50** | 16.4 | 3.82E+08 | + | + | B.1.617.2 |
| **51** | 19.3 | 5.58E+07 | + | - | B.1.617.2 |
| **52** | 20.7 | 2.12E+07 | + | + | B.1.617.2 |
| **53** | 28.1 | 1.55E+05 | - | - | B.1.617.2 |

| | | | | | |
|---|---|---|---|---|---|
| **54** | 26.6 | 4.14E+05 | + | + | B.1.617.2 |
| **55** | 18.7 | 8.02E+07 | + | + | B.1.617.2 |
| **56** | 19.5 | 4.76E+07 | + | + | B.1.617.2 |
| **57** | 33.4 | 4.49E+03 | + | + | B.40 |
| **58** | 26.0 | 6.02E+05 | + | - | B.1.617.2 |
| **59** | 24.5 | 1.63E+06 | + | - | B.1.617.2 |
| **60** | 30.8 | 2.52E+04 | - | + | B.1.617.2 |
| **61** | 19.3 | 5.35E+07 | + | + | B.1.617.2 |
| **62** | 24.0 | 2.34E+06 | + | + | B.1.617.2 |
| **63** | 29.2 | 7.31E+04 | + | + | B.1.617.2 |
| **64** | 22.2 | 8.01E+06 | + | + | N/A |
| **65** | 34.4 | 2.25E+03 | - | - | AY.4 |
| **66** | 17.5 | 1.81E+08 | + | + | B.1.617.2 |
| **67** | 18.1 | 1.20E+08 | + | + | B.1.617.2 |
| **68** | 17.9 | 1.42E+08 | + | + | B.1.617.2 |
| **69** | 23.0 | 4.57E+06 | + | - | B.1.617.2 |
| **70** | 22.6 | 5.91E+06 | + | + | B.1.617.2 |
| **71** | 26.2 | 5.26E+05 | - | - | B.1.617.2 |
| **72** | 23.3 | 3.77E+06 | - | - | B.1.617.2 |
| **73** | 29.4 | 6.50E+04 | - | + | B.1.617.2 |
| **74** | 19.2 | 5.87E+07 | + | + | B.1.617.2 |
| **75** | 16.2 | 4.41E+08 | + | + | B.1.617.2 |
| **76** | 29.6 | 5.67E+04 | - | - | B.1.617.2 |
| **77** | 18.1 | 1.19E+08 | + | + | B.1.617.2 |
| **78** | 17.3 | 2.06E+08 | + | + | B.1.617.2 |

N/A - lineage was not available

**Supplementary Table 7. Comparative RT-PCR cycle threshold values and viral loads between antigen positive and cell culture positive clinical samples.**

| | | Ct value | Viral load* |
|---|---|---|---|
| **Antigen test positive** | Median (95% CI) | 20.00 (19.3 to 23.0) | $8.6x10^{8}$ ($1.1x10^{7}$ to $1.9x10^{8}$) |
| | Maximum | 33.4 | $1.8x10^{8}$ |
| | Minimum | 16.0 | $5.5x10^{3}$ |
| **Cell culture Positive** | Median (95% CI) | 20.3 (19.3 to 23.0) | $7.4x10^{7}$ ($5.8x10^{6}$ to $1.8x10^{8}$) |
| | Maximum | 33.4 | $1.8x10^{8}$ |
| | Minimum | 16.0 | $5.5x10^{3}$ |

* Viral load presented as RNA copies/mL
Ct – Cycle threshold

**Supplementary Figure 1. Logistic regression analysis for antigen test sensitivity.** Rapid antigen tests were compared head-to-head using identical panels of fifteen 2-fold dilutions. Logistic regression plots are shown with mean Ct value (and 95% CI) represented by vertical red lines for all tests where sufficient data points were available for binomial analysis. The remaining tests indicated perfect separation (clear distinction between positive and negative separation) and did not fit a binomial curve. Shown in blue is the 95% CI for the regression curve. Where error bars were too wide, the 95% CI was not plotted.

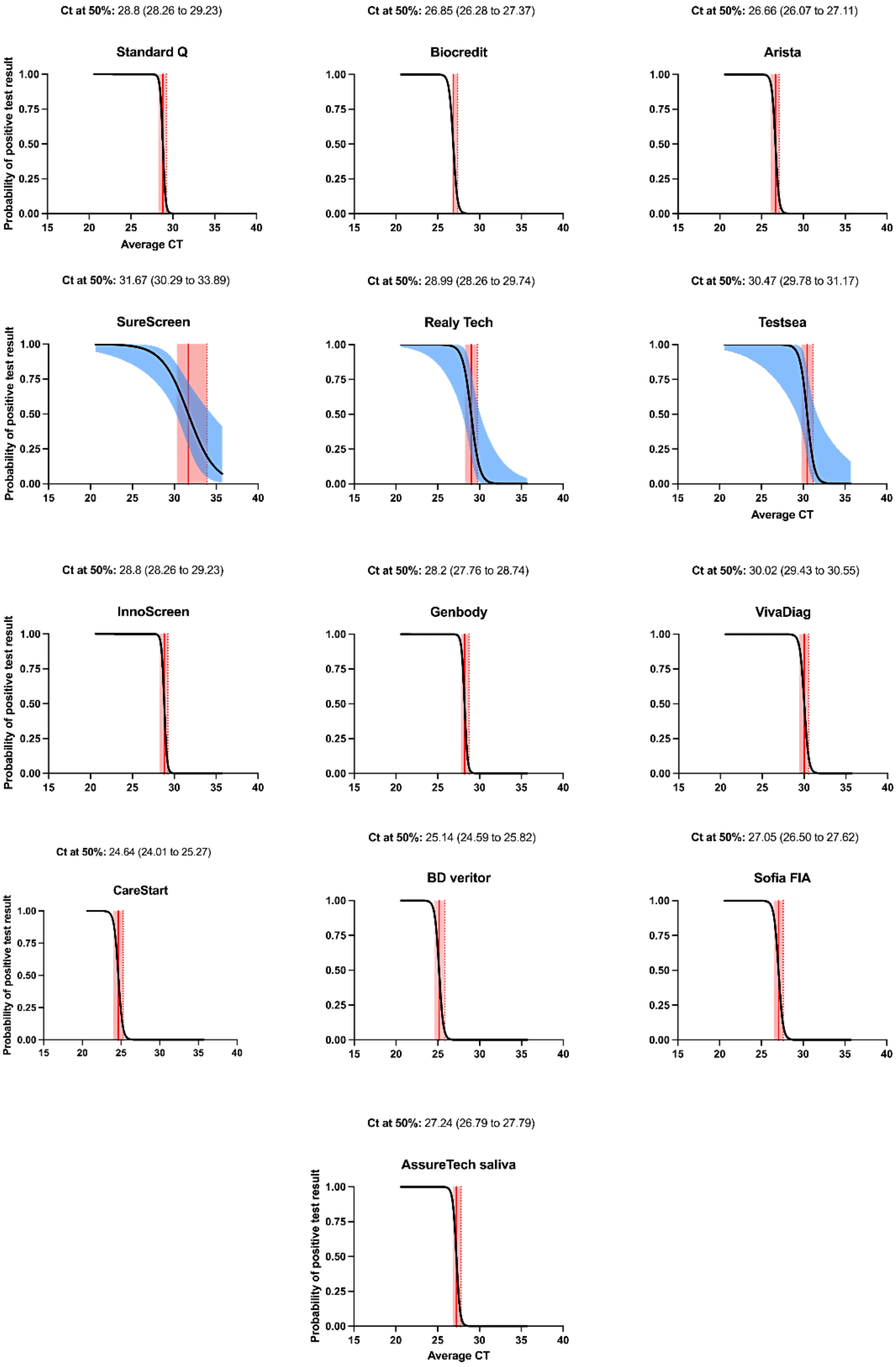
Ct at 50%: 28.8 (28.26 to 29.23)
Standard Q
Ct at 50%: 26.85 (26.28 to 27.37)
Biocredit
Ct at 50%: 26.66 (26.07 to 27.11)
Arista
Ct at 50%: 31.67 (30.29 to 33.89)
SureScreen
Ct at 50%: 28.99 (28.26 to 29.74)
Realy Tech
Ct at 50%: 30.47 (29.78 to 31.17)
Testsea
Ct at 50%: 28.8 (28.26 to 29.23)
InnoScreen
Ct at 50%: 28.2 (27.76 to 28.74)
Genbody
Ct at 50%: 30.02 (29.43 to 30.55)
VivaDiag
Ct at 50%: 24.64 (24.01 to 25.27)
CareStart
Ct at 50%: 25.14 (24.59 to 25.82)
BD veritor
Ct at 50%: 27.05 (26.50 to 27.62)
Sofia FIA
Ct at 50%: 27.24 (26.79 to 27.79)
AssureTech saliva
Probability of positive test result
Average CT

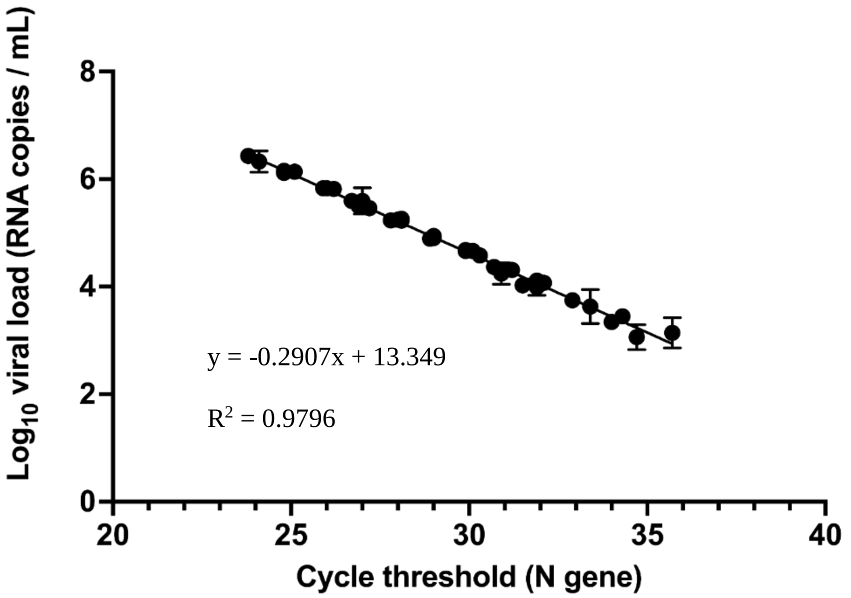


**Supplementary Figure 2. Linear regression analysis of in-house N-gene Ct value against viral load.** The average N-gene Ct value was plotted against quantified SARS-CoV-2 Delta strain (B.1.617.2) from each sample dilution. Correlation coefficient and equation for line of best fit are shown and error bars represent the mean ± SD from n=3 replicates.
